# Impact of Vaccine Failure on the Transmission Dynamics of Measles in Nigeria

**DOI:** 10.1101/2021.02.25.21252459

**Authors:** Ann Nwankwo, Enahoro. Iboi, Daniel Okuonghae

## Abstract

Measles is a vaccine preventable disease. However, it is still a major public health challenge in Nigeria.We therefore formulate a mathematical model for the transmission of measles with a two dose vaccination strategy and weaning of vaccine derived immunity. Using weekly measles cases for Nigeria in 2020 from the Nigeria Center for Disease Control (NCDC), the model was validated. This modelling study via numerical simulations showed that there is a possibility of disease control with a ten fold increase in the vaccination rates. Also, it was shown that primary vaccine failure has more impact on disease dynamics than secondary vaccine failure. Thus control strategies should not just focus on increase the vaccination rates but also look at measures that will help in reducing primary vaccine failure.

## 1 Introduction

Measles is a contagious viral disease. It is a major cause of death (More than 140,000 people died from measles in 2018) among young children globally, despite the availability of a safe and effective vaccine [32]. It was reported in [34] that measles cases increased from 132,490 in 2016 to 869 770 in 2019 globally. This is the highest number reported since 1996 with increases in all WHO regions [34]. Global measles deaths also climbed to nearly a 50% increase since 2016, claiming an estimated 207 500 lives in 2019 alone [34]. Measles is spread via droplets from the nose, mouth or throat of infected persons when they cough or sneeze. Since there is no specific treatment for measles, routine vaccination for children is the best public health strategy for preventing the disease [32]. Initial symptoms usually appear between 10 to 12 days after infection which include high fever, runny nose, bloodshot eyes, tiny white spots on the inside of the mouth and a rash that develops several days later [32].

Measles is endemic in Nigeria, with repeated outbreaks occurring at irregular intervals [13]. Transmission occurs through all months of the year with peaks in the dry season (February, March and April) [12]. Nigeria is one of only ten countries in the world with measles vaccine coverage of less than 50% [30]. Of the estimated 19.8 million infants not vaccinated with at least one dose of measles vaccine through routine immunization in 2019, Nigeria had about 3.3 million unvaccinated infants [34]. Also, quite recently, Nigeria introduced a second dose of measles vaccine into her routine immunization schedule in November 2019 [36] all in a bid to curb the spread of the disease.

Although exposure to the measles is steadily decreasing due to mass vaccination, millions of people are being protected by immunity induced by attenuated vaccines, there are doubts about the quality and duration of vaccine-induced immunity [29, 16]. There are two types of vaccine failure attributed to measles vaccine, primary failure and secondary failure. Primary failure indicates that the vaccine has not taken and fails to induce any immunological response [33]. Measles infection after vaccination is thought to largely arise from primary vaccine failure [33]. The fraction of vaccinees who develop protective antibody levels following measles vaccination depends on a number of factors. Some of which include the presence of maternal antibodies and the immunologic maturity of the vaccinee (low vaccination age), as well as the dose and strain of vaccine virus [2]

However, clinical infection after a prior immune response to vaccination (secondary vaccine failure) has been reported. There has been studies showing evidence of weaning immunity in vaccinated individuals (waning of primary antibody response over time) [20, 38] (and other references therein). Thus, individuals with low antibody titers become at risk of sub-clinical infections [20]. Though most cases of vaccine preventable disease like measles and mumps arise from geographical and social clusters of unvaccinated individuals, an increasing number of reported cases are also seen in sufficiently vaccinated communities [38]. This suggests that immune waning may be an important cause of re-emergence of vaccine preventable diseases (VPDs) [38]. Particularly. Hamami *et al*. (2017) reported that in 2015, 67% of mumps cases in Scotland were fully vaccinated individuals (1 and 2 doses confounded) [11]. Recent evidence suggests that vaccine derived immunity might be less protective than was previously assumed [20] and thus, there is a growing tension that a substantial proportion of vaccinees who respond to a measles vaccine are or will become susceptible to clinical or sub-clinical measles infection [20].

Mathematical models have been used over the years to study the dynamics of diseases. Some of these include the studies in [6, 19, 22, 23, 26]. Thus, many studies have attempted to study the transmission dynamics of measles. Mossong *et al*. [20] using a mathematical model, assessed the epidemiological consequence when vaccinated individuals are able to transmit the measles virus. The model predicted that 80% of all seroconverted vaccinees will have titers below the protective threshold after a long-term absence of circulating virus [20]. Garba *et al*. [9] designed and analyzed a mathematical model, for measles transmission in a population and assessed the combined impact of vaccination and treatment of measles in the population. Aldila and Asrianti [1] analyzed a model of measles with two-step vaccination and quarantine in their work. The model puts into different compartments individuals with one dose of measles vaccine and those with two doses of the vaccine. They used their model to study the impact of quarantine to control the spread of measles compared to the vaccination strategy. The study in [7] constructed both a determinstic and a stochastic model for measles transmission involving vaccination and a hospitalized compartment to investigate the most influential parameters for proposing a control strategy.They proposed that providing treatment accesses is better than vaccinating when an outbreak occurs [7]. Also, Memon *et al*. (2020) reported that high vaccine coverage rate is vital for disease control [17]. Again, the study in [37] analyzed a dynamic model incorporating both periodic transmission and asymptomatic infection with waning immunity with the aim of identifying parameters that influence seasonal fluctuation of measles as well as provide optimal control measures to minimize the number of infected individuals and costs.

Therefore, the objective of this study is to glean, via a mathematical model, a public health control strategy based on vaccination for effective management and possible elimination of measles in Nigeria. This will be done by assessing the role of both primary and secondary vaccine failures. The paper is organized as follows. The model is formulation is given in Section 2 and the model is analyzed in Section 3. Model validation is carried out in section 4 while Numerical simulations are considered in Section 5. The findings from this study are summarized in Section 6.

## 2 Model Formulation

We consider a deterministic compartmental model in a population subject to routine vaccination program. The total population given as *N*_*h*_(*t*) is divided into the sub-populations of susceptibles (*S*(*t*)), single dose vaccinees who did not seroconvert (*V*_1_(*t*)) single dose vaccinees who seroconverted (*V*_2_(*t*)), two dose vaccinees who did not seroconvert (*V*_3_(*t*)), two dose vaccinees who seroconverted with temporary immunity (poor responders) (*V*_4_(*t*)), vaccinees with weaned immunity (*V*_5_(*t*)) unvaccinated exposed individuals(*E*_1_(*t*)), vaccinated exposed individuals(*E*_2_(*t*)), unvaccinated infectious individuals (*I*_1_(*t*)), vaccinated infectious (non-clinical infection) individuals (*I*_2_(*t*)), and recovered individuals (*R*(*t*)) at time t, so that

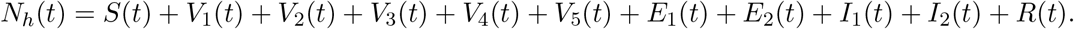

Susceptibles enter the population at a constant rate Π. This population is depleted following measles infection (*λ*_*m*_), first dose vaccination (*α*_1_) and natural mortality (*µ*). So, we have

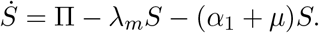

A proportion *p* of susceptibles who receive the first dose of the measles vaccine do not seroconvert and thus join the class *V*_1_. The remaining proportion join the *V*_2_ class as studies have shown that measles epidemics can also occur in highly vaccinated populations due to a variety of factors which include failure to seroconvert and waning of vaccine-induced immunity [20] (and other references therein). Individuals in the *V*_1_ class can either die naturally, get infected or receive a second dose of the measles vaccine (*α*_2_). While individuals in the *V*_2_ class can lose their immunity (*ϕ*_1_), die naturally or receive a second dose of the measles vaccine at the rate *α*_3_. These give

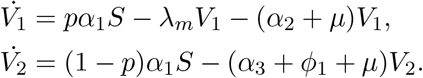

A fraction *f* of vaccinees in the *V*_1_ may fail to respond to the vaccine and so join the *V*_3_ class and are considered susceptible to the measles infection (*λ*_*m*_). Another fraction *q* develop temporary immunity and enter the *V*_4_ class whose immunity weans at a rate *ϕ*_2_. The remaining fraction 1 − *f* − *q* develop strong immune response and enter the *R* class. Again, a fraction of vaccinees *r* from the *V*_2_ class develop temporary immunty and so join the *V*_4_ class and the remaining fraction 1 − *r* join the *R* class. Thus, we have

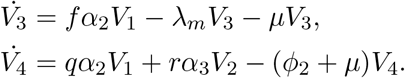

The class *V*_5_ of vaccinees with weaned immunity is generated by vaccinees from *V*_2_ and *V*_4_ classes at the rates *ϕ*_1_ and *ϕ*_2_. This population is decreased by infection at a reduced rate *ϵλ*_*m*_ (where *ϵ* shows the effect of immunological memory) and natural death. Thus, we have

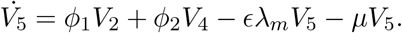

Individuals in the *E*_1_ class is generated by the infection of those in the *S, V*_1_ and *V*_3_ classes. This population will be decreased by the development of symptoms at the rate *γ*_1_ and by natural mortality.Similarly, individuals in the *E*_2_ class is generated by the infection of those in the *V*_5_ class. This population will be decreased by the development of symptoms at the rate *γ*_2_ and by natural mortality. Thus, we have

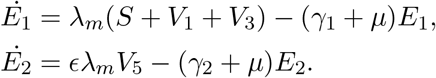

Individuals in the *I*_1_ class is generated by the development of symptoms of unvaccinated exposed individuals and by the proportion of vaccinated infectious individuals who develop symptoms (*hγ*_2_). This class is also decreased by recovery at the rate *σ*_1_ and by natural mortality.Similarly, individuals in the *I*_2_ class is generated by the progression of those in the *E*_2_ class. This population will be decreased by recovery at the rate *σ*_2_ and by natural mortality. Thus, we have

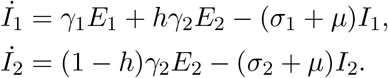

Finally, the population of recovered individuals is generated by fraction of those who developed permanent immunity following a second dose of measles vaccine from the *V*_1_ and *V*_2_ classes as well as recovery from symptomatic and asymptomatic infections. The population is decreased by natural mortality. Thus, we have

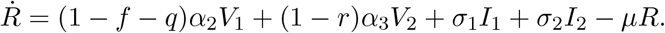

The model is therefore given by the following deterministic, system of nonlinear ordinary differential equations:

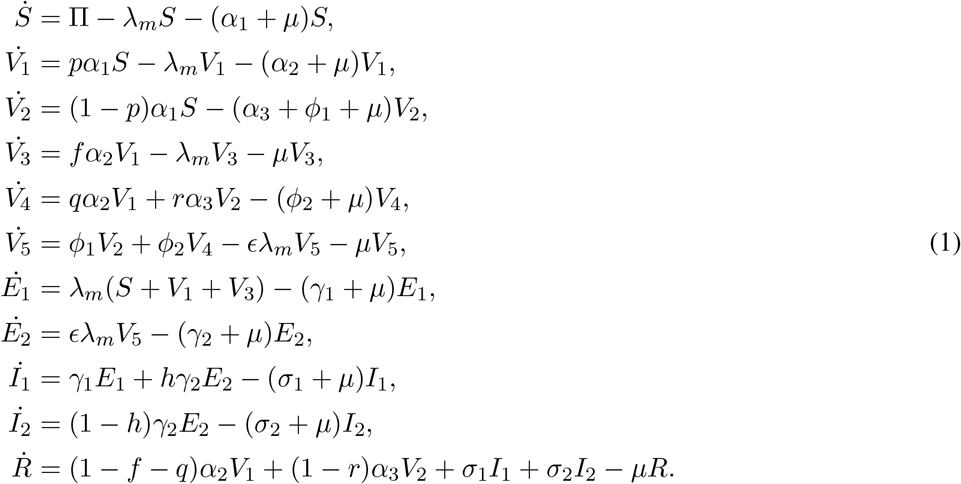

The rate of infection of the model (1) are given as 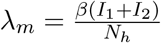.

To accounts for the reduced infectiousness of vaccinated infectious individuals (*I*_2_) we assumed that *σ*_2_ *> σ*_1_ (ie, the average duration of infectiousness of vaccinated individuals is assumed to be less than that of unvaccinated individuals). This reduced infectiousness is based on the observation that symptoms in vaccinated individuals if they occur at all, generally tend to be milder and of shorter duration than those in unvaccinated individuals [20]. We also assumed that the fraction of vaccinees who develop permanent immunity to measles do so after the second dose of the measles vaccine.

The flow diagram of the model (1) is given in Figure 1 while the meanings of the variables and parameters of the model (1) are given in Table 1.

**Table 1:** Description of variables and parameters in the model (1).

| **Variable** | **Interpretation** |
| --- | --- |
| $S$ | Population of susceptible individuals |
| $V_1$ | Vaccinated individuals with one dose who did not seroconvert |
| $V_2$ | Vaccinated individuals with one dose who seroconverted |
| $V_3$ | Vaccinated individuals with two doses who did not seroconvert |
| $V_4$ | Vaccinated individuals with two doses who seroconverted (temporarily immuned) |
| $V_5$ | Population of vaccinated individuals with waned immunity |
| $E_1$ | Non vaccinated exposed individuals |
| $E_2$ | Vaccinated exposed individuals |
| $I_1$ | Non vaccinated infectious individuals |
| $I_2$ | Vaccinated infectious individuals |
| $R$ | Recovered individuals |
| **Parameter** | **Interpretation** |
| $\Pi$ | Recruitment rate |
| $\beta$ | Effective contact rate |
| $\alpha_1$ | Vaccination rate for first dose |
| $\alpha_2$ | Vaccination rate for second dose for those in $V_1$ class |
| $\alpha_3$ | Vaccination rate for second dose for those in $V_2$ class |
| $p$ | Fraction of vaccinated with one dose who did not seroconvert |
| $f$ | Fraction of vaccinated with two doses who did not seroconvert |
| $q$ | Fraction of vaccinated from $V_1$ with two dose who seroconverted (temporary immunity) |
| $r$ | Fraction of vaccinated from $V_2$ class with two dose who seroconverted (temporary immunity) |
| $\epsilon$ | Reduced susceptibility of individuals with waned immunity |
| $\gamma_1, \gamma_2$ | Progression rate from exposed to infectious |
| $\sigma_1, \sigma_2$ | Recovery rate |
| $\phi_1, \phi_2$ | Weaning rate of vaccine derived immunity |

**Figure 1:**
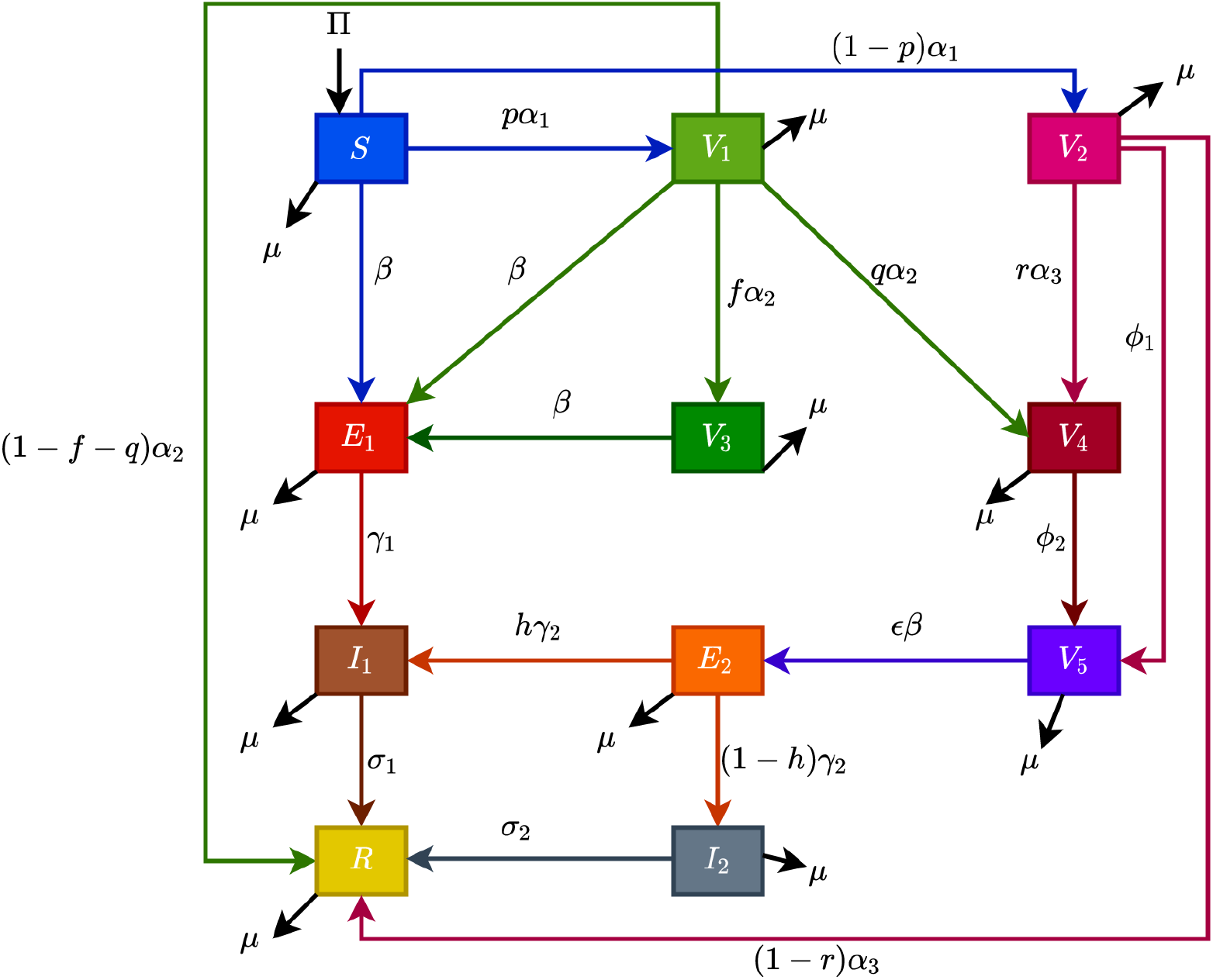
Schematic diagram of the model (1).

## 3 Analysis of the model

The qualitative properties of the model (1) will be explored in this section.

### 3.1 Basic properties

The variables and parameters of the model (1) are assumed to be non-negative for *t* ≥ 0 since they are different human statues. It can be shown that the feasible region 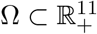 defined as

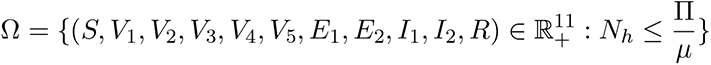

is positively invariant for the model (1).

#### Theorem 3.1.

*The closed set* Ω *is positively invariant for the model* (1).

#### Proof.

Given that the total population *N*_*h*_ = *S* + *V*_1_ + *V*_2_ + *V*_3_ + *V*_4_ + *V*_5_ + *ϵ*_1_ + *ϵ*_2_ + *I*_1_ + *I*_2_ + *R*. The rate of change of the total population is given as 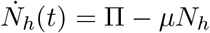. The standard comparison theorem can be used to show that

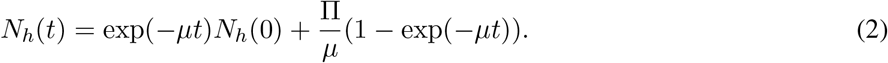

Particularly, 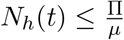 if 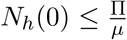. thus, all solutions of the model (1) with initial conditions in Ω will remain in Ω. Hence, the region Ω is a positively invariant region of the model (1).

### 3.2 Local asymptotic stability of disease free equilibrium

The control reproduction number associated with the model (1) is computed using the approach in [4, 5]. We Linearize the model (1) at the disease-free state

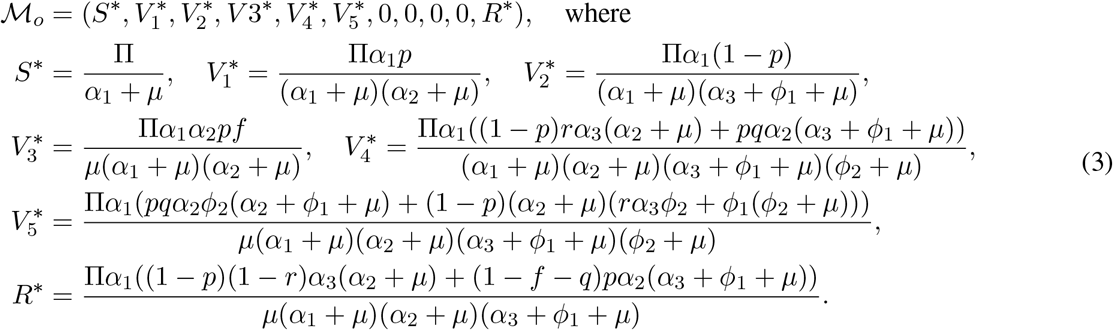

The *F* (matrix of the new infection terms) and *V* (the M-Matrix) of the remaining transfer terms, associated with the model (1) are given below.

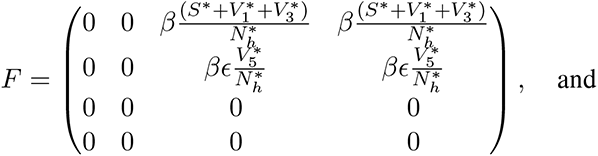

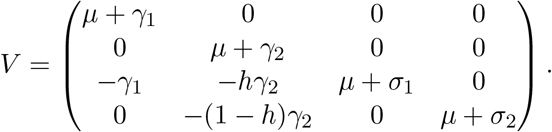

The control reproduction number ℛ_*v*_ is then given as the spectral radius as *ρ*(*FV* ^−1^) which is written as.

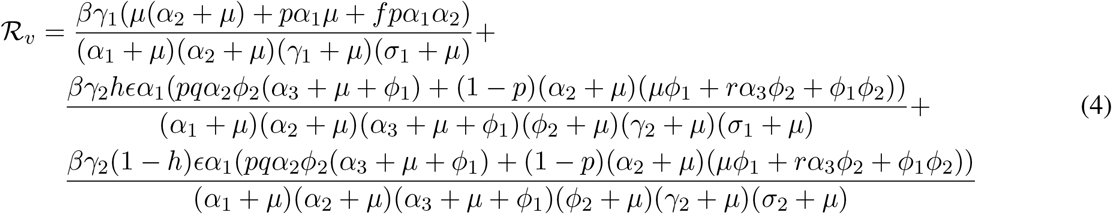

The result below follows from Theorem 2 in [5].

#### Theorem 3.2.

*The disease free state* (ℳ_*o*_) *of the model* (1) *is locally asymptotically stable if* ℛ_*v*_ *<* 1 *and unstable if* ℛ_*v*_ *>* 1.

By Theorem 3.2, biologically speaking, measles can be controlled in Nigeria, when _*v*_ *<* 1 and the initial conditions are in the region of attraction of ℳ_*o*_.

The quantity ℛ_*v*_ is the control reproduction number of the model (1). It is the sum of constituent reproduction numbers associated with the number of new infections generated by symptomatic and asymptomatic infectious individuals. The first term is the number of new cases of infections generated by unvaccinated symptomatic infectious individuals. It is the product of the transmission probability 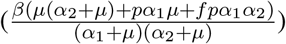, the probability that they survived the exposed (*E*_1_) class 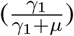 and the average time they spend in the *I*_1_ class 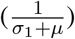. The second term is the number of new cases of infections generated by vaccinated symptomatic infectious individuals. It is the product of the transmission probability 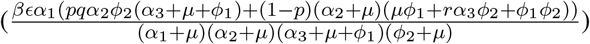, the probability that they survived the exposed (*E*_2_) class 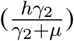 and the average time they spend in the *I*_1_ class 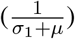. The third term is the number of new cases of infections generated by vaccinated asymptomatic infectious individuals. It is the product of the transmission probability 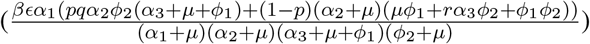, the probability that they survived the exposed (*E;*_2_) class 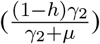 and the average time they spend in the *I*_2_ class 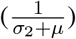.

### 3.3 Global asymptotic stability of disease free state (ℳ_0_)

The global stability of the disease-free state of the model (1) will be established. We claim the following result,

#### Theorem 3.3.

*The disease-free state of the model* (1) *is GAS whenever* ℛ_*v*_ *<* 1

The proof of Theorem 3.3 is shown in Appendix A.

The biological significance of Theorem 3.3 is that measles can be effectively controlled (if not eliminated) if the control reproduction number ℛ_*v*_ would not only be brought to but also maintained at a value less than unity.

### 3.4 Existence of disease endemic state

Let the disease endemic state of the model (1) be denoted by

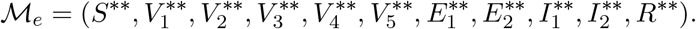

Solving the equations of the model (1) in terms of the force of infection at steady state, we have that

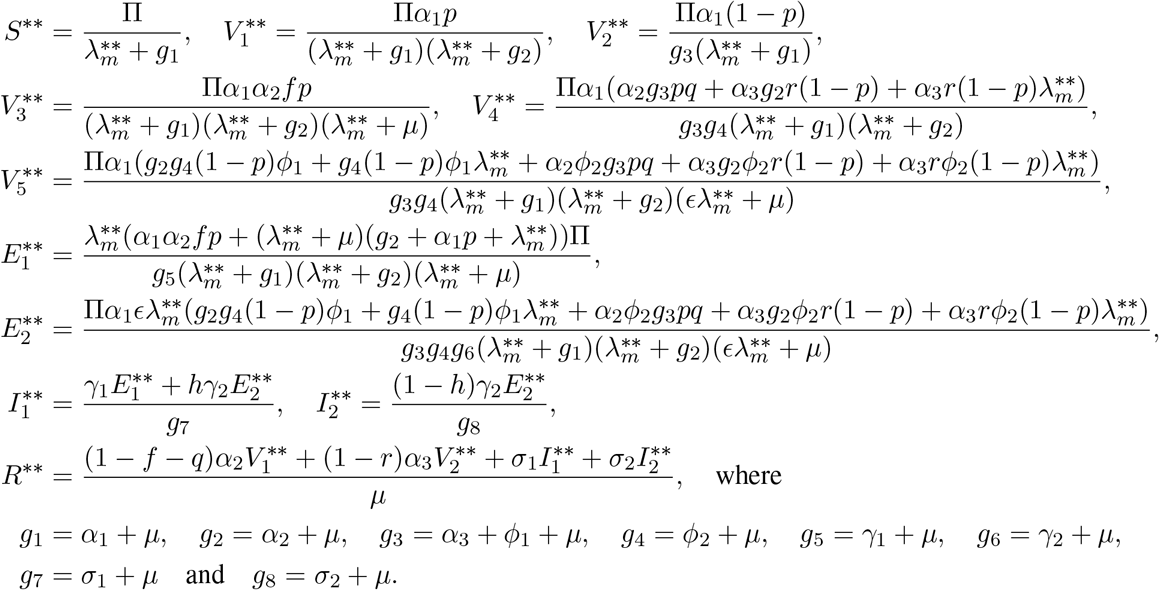

Since 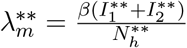, substituting 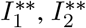, and 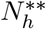 we have that

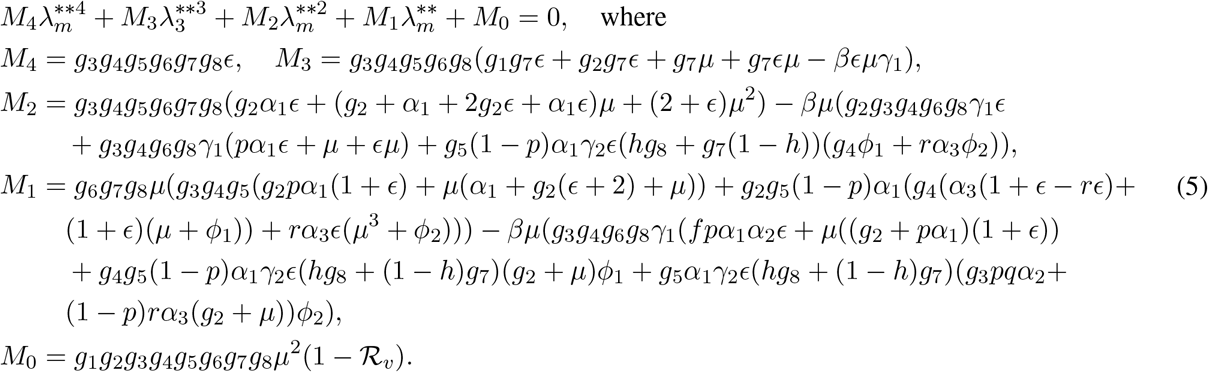

It is clear to see that *M*_4_ is positive from the expressions above. Also, *M*_0_ *<* 0 if ℛ_*v*_ *>* 1. Thus, using the descrates rule of signs, the polynomial (5) will have at least one positive root. Therefore, the following result is established.

#### Theorem 3.4.

*The model* (1) *has at least one positive equilibrium whenever* ℛ_*v*_ *>* 1.

### 3.5 Global asymptotic stability of disease endemic state (ℳ_*e*_)

Consider the model (1) with *h* = 0. The global stability of the disease endemic state will now be studied when ℛ_*v*_|_*h*=0_ *>* 1 (that is, when the stability of the disease-free state is violated). We claim the following result

#### Theorem 3.5.

*Suppose that h* = 0 *and* ℛ_*v*_ | _*h*=0_ *>* 1 *for all t* ≥ 0, *then the disease endemic state of model* (1) *is globally asymptotically stable*.

The proof of Theorem 3.5 is given in Appendix B.

The epidemiological significance of Theorem 3.5 is that whenever ℛ_*v*_ | _*h*=0_ *>* 1, for all *t* ≥0, the disease will persist in the population because the infected classes of the model will persist above a certain threshold value at steady state.

## 4 Model fitting and parameter estimation

One important step to be taken during model validation is the use of data to estimate the values of some unknown parameters in the model under consideration. Therefore, weekly measles incidence data was for Nigeria (see Figure 2) was obtained from the Nigeria Center for Disease Control (NCDC) for model validation and also to obtain best fitted values of some unknown biological parameters that are present in the model. The model fitting was carried out using a genetic algorithm (GA) [18] for our function optimizer, implemented in MATLAB; the GA algorithm helps in finding the correct basin of attraction, which then provides the starting values (for the parameters being estimated) for use in the lsqnonlin function in the Optimization Toolbox of MATLAB. Here, we fitted the model using the seven week moving average of the cumulative measles data (obtained from the weekly measles data from the first week to the last week in 2020). The results depicted in Figure 3, shows the fitting of the cumulative measles cases for Nigeria for the period considered. The ranges and baseline values of the parameters of the model are are given in Table 2.

**Table 2:**
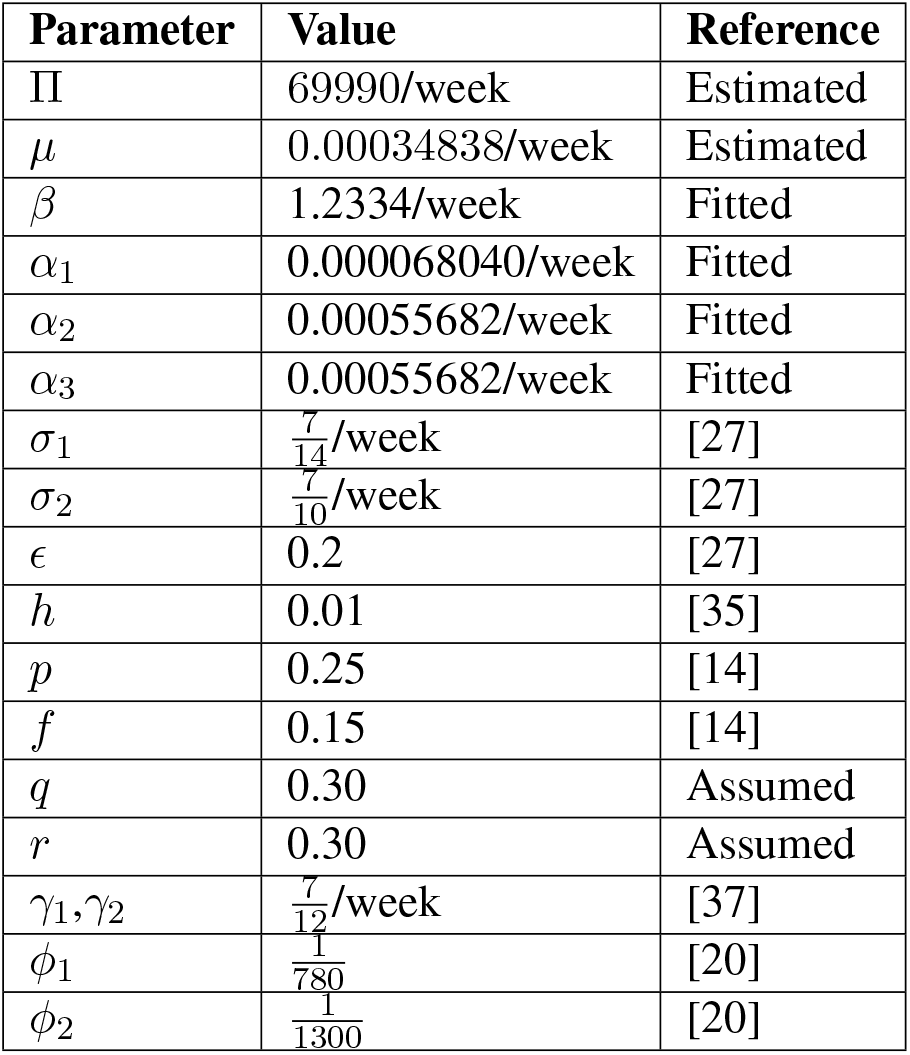
Values of the parameters in the model (1).

**Figure 2:**
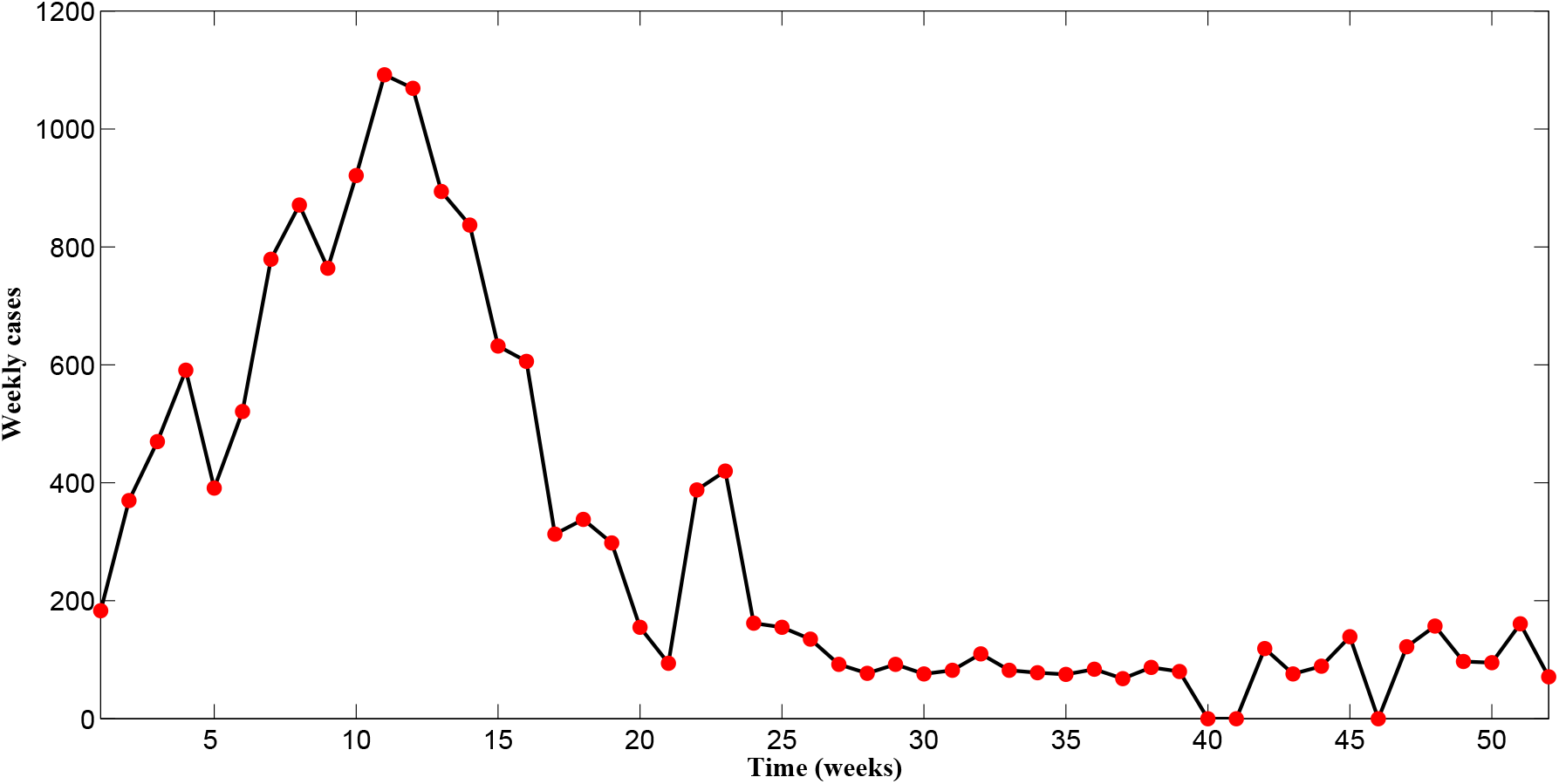
Weekly data of measles incidence in Nigeria from January 2020 to December 2020.

**Figure 3:**
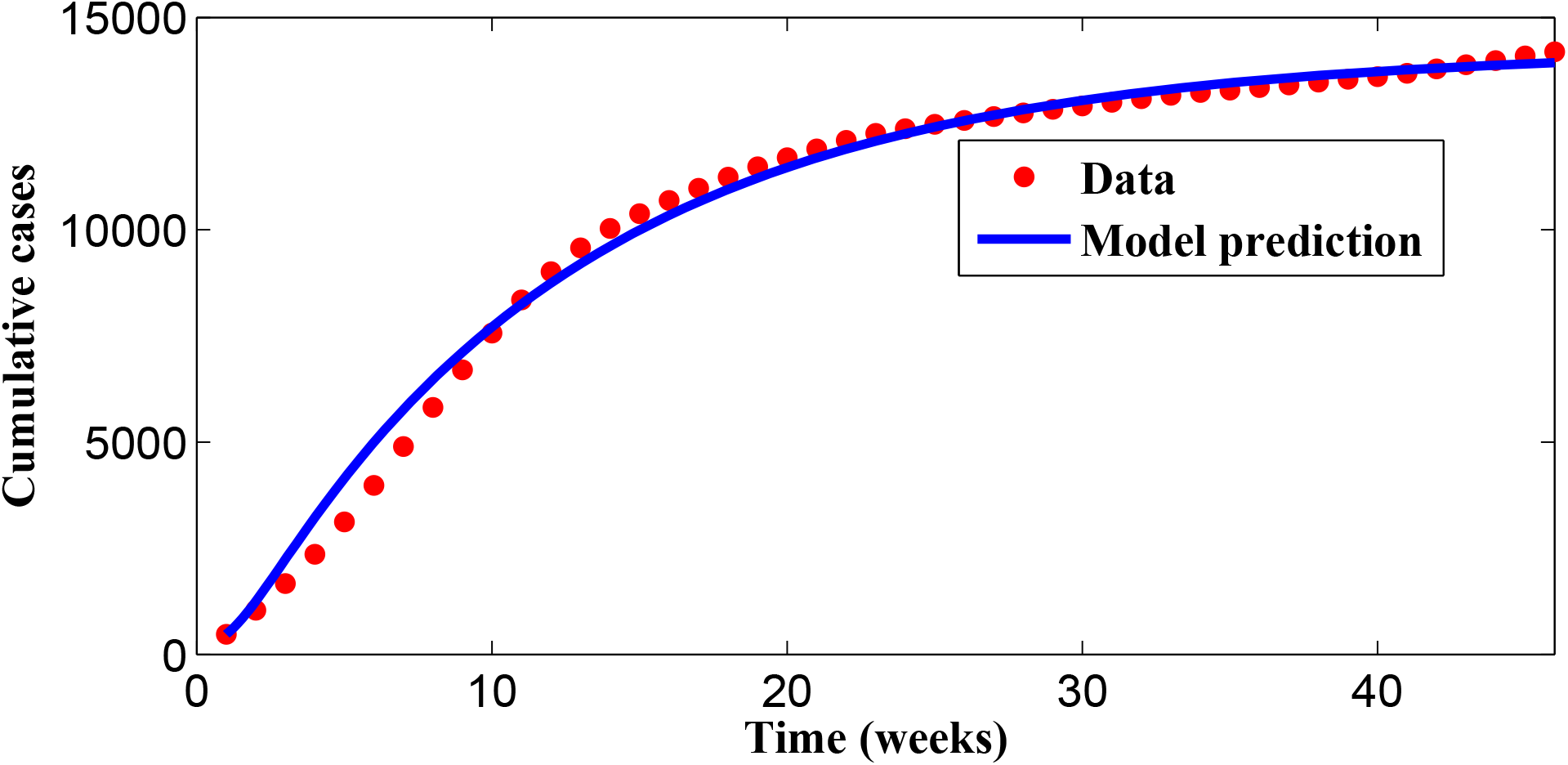
Fitting the cumulative incidence.

## 5 Numerical Simulations

The model (1) is now simulated to illustrate the population-level impact of the vaccination rate and vaccine failure on the transmission dynamics of measles in Nigeria. We investigate the relationship between the effective reproduction number (ℛ _*v*_) of the model (1) and some key parameters of the model (1). Figure 4 is a contour plot of ℛ_*v*_ as a function of the fraction or proportion of vaccinees that did not seroconvert after the first and second doses of the measles vaccine. It is seen from Figure 4(a) that for the present parameter values, it might be impossible to control the infection in the population. This might be due to the very low vaccination rates for both the first and second doses of the measles vaccine. However, with a ten fold increase in the vaccination rates *α*_1_, *α*_2_ and *α*_3_, Figure 4(b) shows the possibility of controlling measles infections in the population.

**Figure 4:**
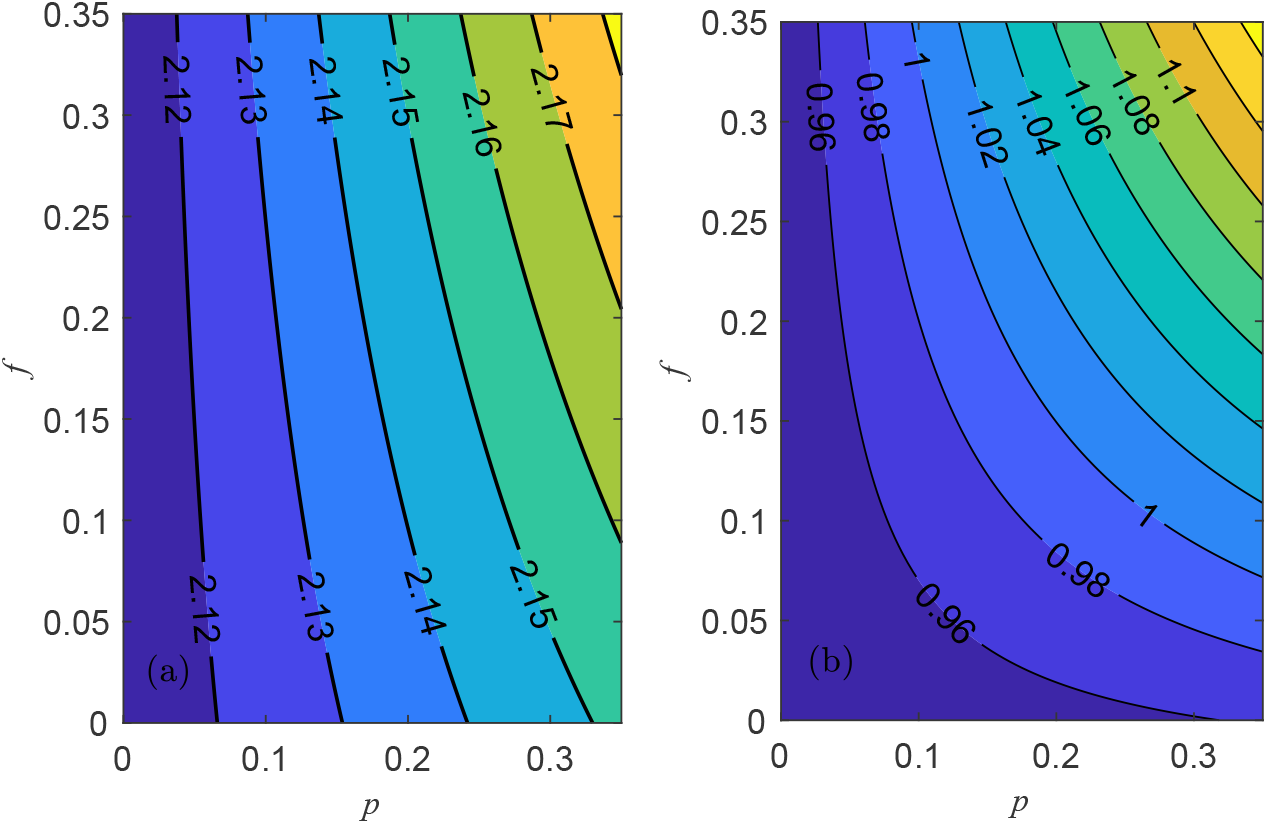
Simulation of the control reproduction number ℛ_*v*_ of the model (1). (a) vaccination rates at baseline (b) ten fold increase in vaccination rates. Other parameter values are as given in Table 2.

The model (1) is solved numerically using parameter values in Table 2 to assess the impact of vaccination as well as primary and secondary vaccine failure on the transmission dynamics of measles in the population. Figure 5 shows the cumulative measles cases and the prevalence of measles in the population if 25% of first dose vaccinees and 15% of second dose vaccinees do not seroconvert upon vaccination in the presence of weaning immunity. Again, we see that for the present parameter values, measles will persist in the population. It is important to state that with the present vaccination level in Nigeria, there will be periodic epidemic cycles (Figure 5(b)). However, the subsequent ones are of lesser magnitude than the previous. These cycles in Figure 5(b) corresponds to the step function curve in Figure 5(a). This is so because after an epidemic, the immunity of the people is boosted; thus, to support another epidemic, a certain amount of time is required for the population to become suffiently susceptible. Therefore, if Nigeria continues to vaccinate at the rate in this study, we may not be able to eliminate measles. However, if vaccination rates are increased, they may see a dratic reduction in the cumulative cases as depicted in Figure 6 where the model was simulated for three different vaccination levels. We see that increasing the vaccination rates (*α*_1_, *α*_2_ and *α*_3_) leads to significant decrease in the cumulative incidence of measles in the population. Showing that vaccination plays a significant role in the dynamics of measles in Nigeria.

**Figure 5:**
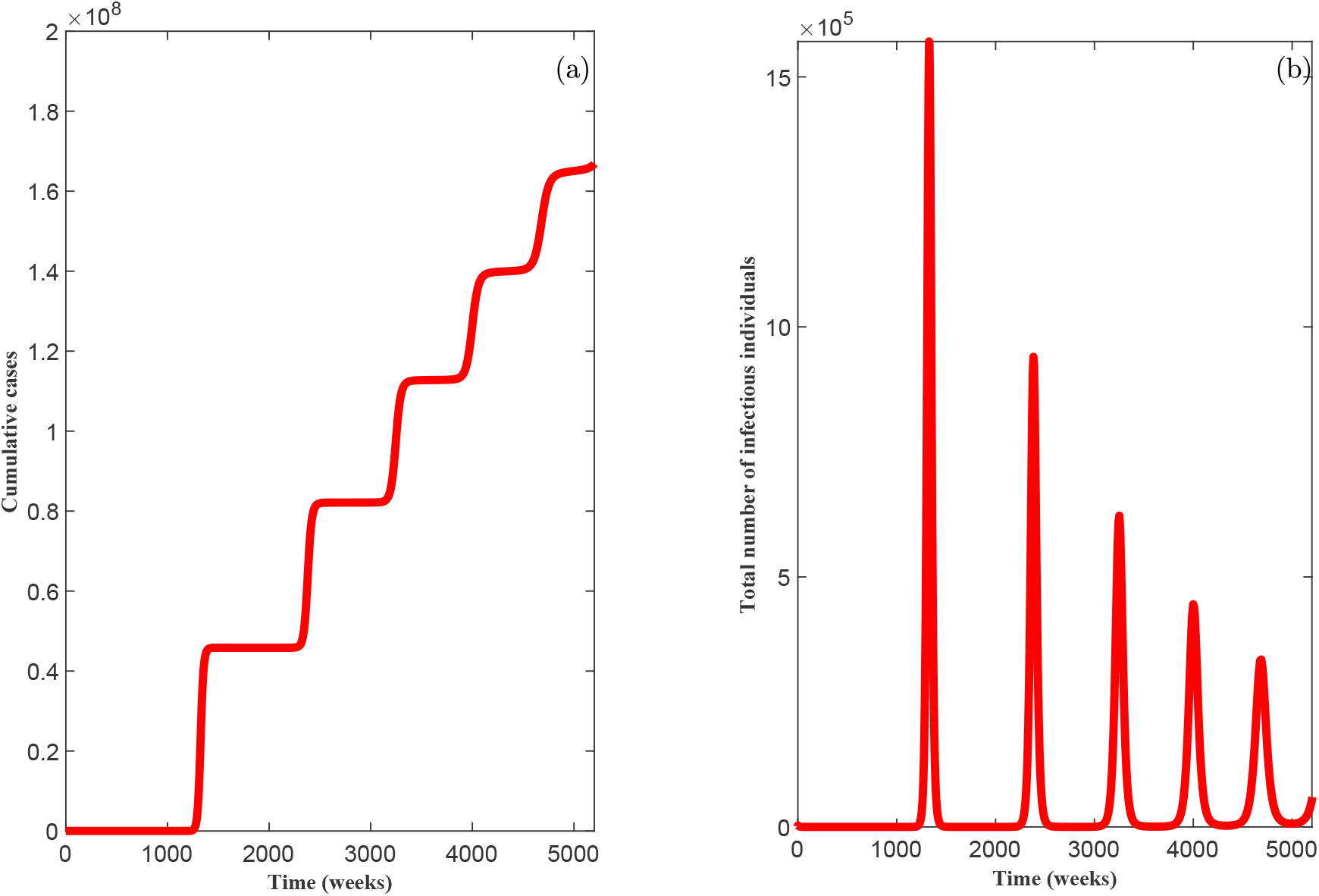
Simulations of the model (1) showing (a) the cumulative incidence of measles (b) prevalence of measles. Parameter values are as given in Table 2

**Figure 6:**
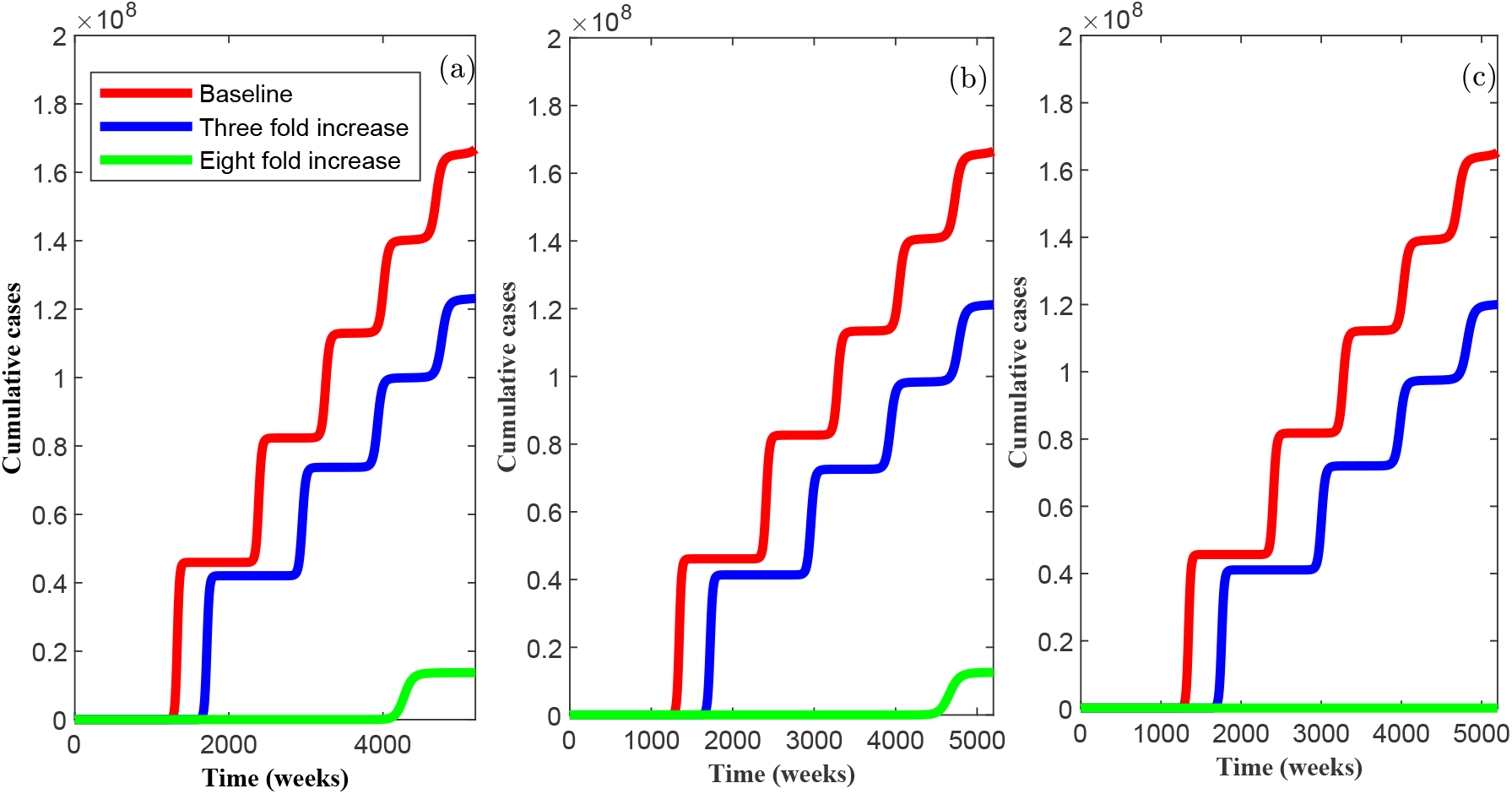
Simulations of the model (1) showing the cumulative incidence with varying values of the vaccination rates. (a) Other parameters at baseline (b) 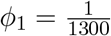 and 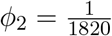 (c) *p* = 0.15 and *f* = 0.05.

Comparing Figures 6 (a),(b) and (c) we can see that primary vaccine failure has more impact than secondary vaccine failure on the dynamics of measles transmission in Nigeria. More specifically, results from Figure 6(b) show that even with an eight fold increase in the vaccination rates, increasing the duration of immunity of first dose vaccinees from 15 to 25 years and second dose vaccinees from 25 to 35 years, it might still be impossible to control measles in the population. However, results from Figure 6(c) shows that measles can be controlled if there is an eight fold increase in the vaccination rates and with an 85% seroconversion rate of first dose vaccinees and 95% seroconversion rate of second dose vaccinnees

In particular, Figures 6(b) and 6(c) show that the impact of primary and secondary vaccine failure is more evident, with an increase in the vaccination rates. This result is in line with reports from [10] which revealed that that the effects of subclinical infections resulting from secondary vaccine failure is more significant when immunization rate is high. This according to the study in [10] is due to the fact that increase in vaccination leads to reduction of natural boosters of immunity which will have a further impact on the rate of weaning of immunity (secondary vaccine failure). In addition, Figure 6(b) reveals that although most cases of measles still come from unvaccinated individuals which includes those with a primary vaccine failure, secondary vaccine failure may still contribute to the incidence of measles in the population. This is so because an increase in the duration of vaccine induced immunity of *ϕ*_1_ from 15 to 25 years and *ϕ*_2_ from 25 to 35 years lead to 8.45% decrease in the cumulative incidence of measles when there is an eightfold increase in the vaccination rates.

## 6 Discussions and Conclusion

A mathematical model for the transmission dynamics of measles is formulated (and rigorously analyzed) and used to assess the impact of primary and secondary vaccine failure on the transmission dynamics of measles. Some of the theoretical findings are given below.

The measles free equilibrium is shown to be both locally and globally asymptotically stable whenever the associated control reproduction number ℛ_*v*_ is less than unity. However, in the case where the associated control reproduction number is greater than unity (ℛ_*v*_ *>* 1), the model (1) has at least an endemic equilibrium which makes for the persistence of the disease.

Numerical simulations show that vaccination plays a great role in the transmission dynamics of measles in Nigeria as increasing the vaccination rates led to significant reduction in measles cumulative cases. Also, primary vaccine failure was shown to have more meaningful impact on the transmission dynamics of measles in Nigeria than secondary vaccine failure. In Nigeria, studies have shown that the proportion of vaccinees who do not seroconvert resulting in a primary vaccine failure (which may be due to a number of factor which might include inappropriate practice of the cold chain management, the potency of the vaccine administered and presence of maternal antobodies in vaccinees) is a very important factor in the control of measles in Nigeria [24]. In particular, the study in [24] showed that only 51.3% of primary health care workers has appropriate vaccine handling practice while [25] reported that 73.9% had appropriate vaccine handling practice. Also, [3] reported that 73% of health workers in Oyo state were aware of vaccine handling and storage guidelines with 68.4% having ever read such guidelines. Only 15.3% read a guideline less than 1 month prior to the study. About 43.0% had good knowledge of vaccine management, while 66.1% had good vaccine management practices.

An efficient cold chain vaccine management requires three major elements [25]. These include well trained personnel, reliable transport/storage equipment and efficient management procedures [25](and other references therein) and absence of any of these leads to a deficient cold chain system. Health workers play an important role in maintaining an undisrupted cold chain as they are the last point of contact between the vaccines and the recipient. Hence, it is very pertinent that they be trained and supervised regularly in order to ensure efficient practice of cold chain management [3, 25]. Improper handling has been identified as one of the major reasons for the decline in vaccine potency at the time of vaccine administration [3]. Loss of potency becomes evident when immunised individuals contract the diseases the vaccines were meant to prevent.The study in [8] reported 68.6% seroconversion rate among vaccinees in Ilorin, Kwara State Nigeria [8]. This study also reported that the low seroconvrsion rate is partly due to low vaccine potency as only 50% of the vaccine vials had virus titers above the cut off point recommended by the World Health Organization for measles vaccines.

Again, numerical simulations show that secondary vaccine failure may still to contribute to measles incidence with increase in the vaccination rates. This result is in agreement with the findings in [10, 38]. The study in [10] reported that increase in vaccination leads to reduction of natural boosters of immunity which have a further impact on the rate of weaning of immunity leading to a acumulation of susceptible people. Also, Yang *et al*. (2020), revealed that waning immunity may still have potential epidemiological consequences (with increased vaccination coverage) beyond disease transmission [38]. For example, faster decay of maternal antibodies in newborns of women of childbearing age with vaccine induced immunity (who may be vaccinated at infancy and as such may have little antibody to transfer to their infant) may also lead accumulation of infants who are susceptible before the currently recommended schedule for the first dose of MMR vaccine [35, 38]. Thus leading to an outbreak of measles. Thus, Yang *et al*. (2020), suggested that understanding the interaction between vaccination, weaning and boosting will be important in coming up with optimal control strategies.

In conclusion, this study has emphasized the importance of increasing vaccination coverage as well as managing primary vaccine failure in Nigeria since both have significant impact on measles transmission. The public health implication of these results is that measles can be controlled if protection of susceptible individuals is encouraged by ensuring improved immunization strategies which can be achieved through increased vaccination coverage as well as reducing primary vaccine failure.

## Data Availability

NA

## Appendix A Proof of Theorem 3.3

Consider the model (1) with ℛ_*v*_ *<* 1 for all *t* ≥ 0. Now consider the following Lyapunov function

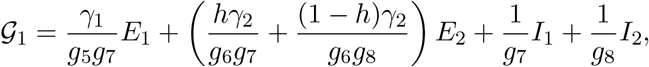

with derivative

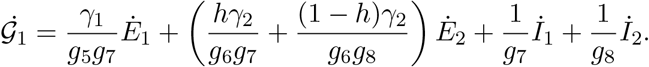

Substistuting the expressions for equations 7 to 10 of model (1) into the above expression, we have that

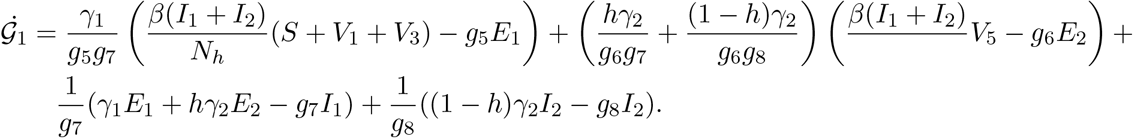

It follows after some algebraic simplification that

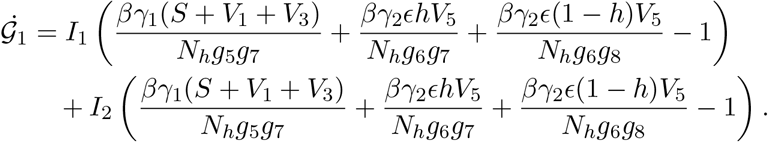

Since 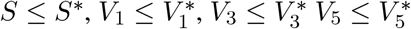 and 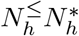, it follows that

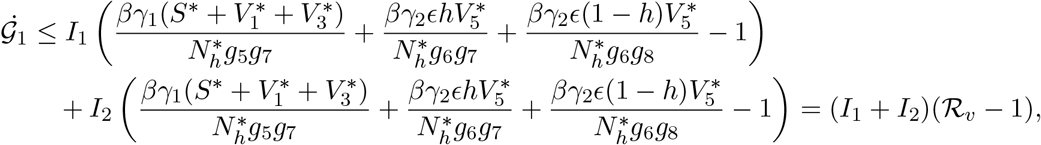

with equality only at ℳ_*o*_. For ℛ_*v*_ *≤* 1, 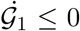 with equality if and only if *E*_1_ = *E*_2_ = *I*_1_ = *I*_2_ = 0. Therefore, 𝒢_1_ is a Lyapunov function in Ω, and it follows from LaSalle’s invariance principle [15] that every solution to the equations in (1) with initial conditions in Ω converges to ℳ_*o*_ as *t → ∞*.

## Appendix B: Proof of Theorem 3.5

*Proof*. Consider the model (1) with *h* = 0 and ℛ_*v*_ *>* 1, so that the associated unique endemic equilibrium exists. Also, consider the non-linear Lyapunov function of the Goh-Volterra type

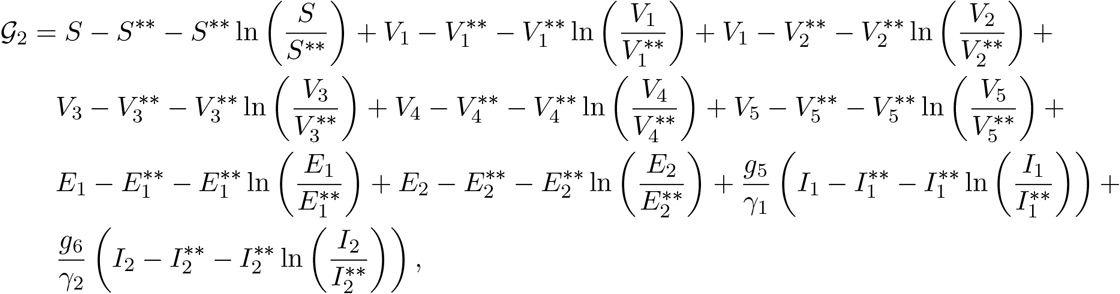

with Lyapunov derivative,

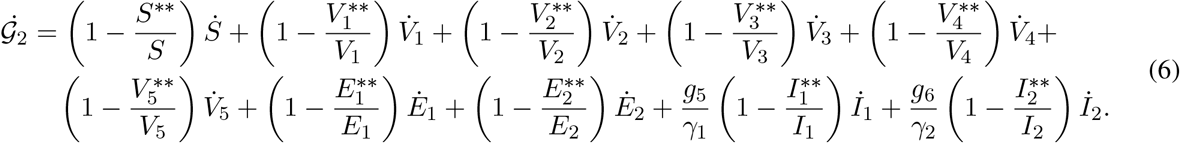

It can be shown from model (1) with *h* = 0, that, at steady state,

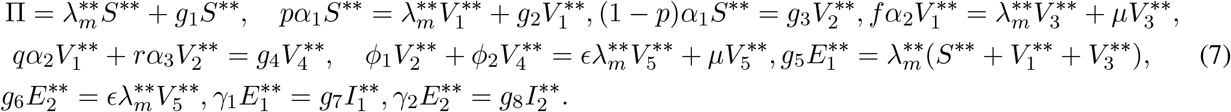

Substituting the time derivatives of *S, V*_1_, *V*_2_, *V*_3_, *V*_4_, *V*_5_, *E*_1_, *E*_2_, *I*_1_ and *I*_2_, in (1) as well as the relations in (7), into (6), we have that (after several algebraic calculations)

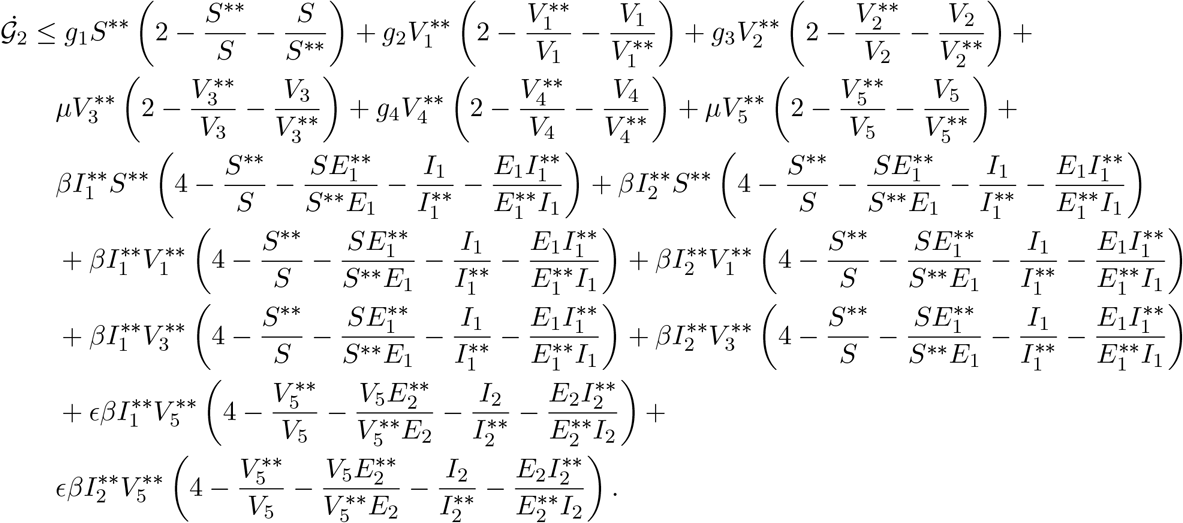

Finally, since, the arithmetic mean is greater than the geometric mean, the following inequalities hold.

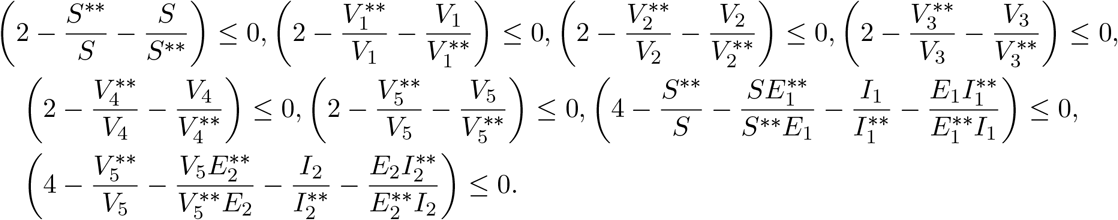

Thus we have that 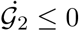 for ℛ_*v*_ *>* 1. Since the relevant variables in the equation for *R* are at endemic steady state, it follows that these can be substituted into the equation for *R* so that *R* → *R*^*\*\**^ as *t → ∞*. Hence, 𝒢_2_ is a Lypapunov function.

